# Benchmarking clinical risk prediction algorithms with ensemble machine learning: An illustration of the superlearner algorithm for the non-invasive diagnosis of liver fibrosis in non-alcoholic fatty liver disease

**DOI:** 10.1101/2023.08.02.23293569

**Authors:** Vivek Charu, Jane W. Liang, Ajitha Mannalithara, Allison Kwong, Lu Tian, W. Ray Kim

**Affiliations:** Quantitative Sciences Unit, Department of Medicine, Stanford University School of Medicine, Stanford, CA; Department of Pathology, Stanford University School of Medicine, Stanford, CA; Division of Gastroenterology and Hepatology, Department of Medicine, Stanford University School of Medicine, Stanford, CA; Department of Biomedical Data Sciences, Stanford University School of Medicine, Stanford CA

**Keywords:** Clinical risk prediction, non-invasive testing, liver fibrosis, machine learning

## Abstract

**Background and Aims:** Ensemble machine learning (ML) methods can combine many individual models into a single ‘super’ model using an optimal weighted combination. Here we demonstrate how an underutilized ensemble model, the superlearner, can be used as a benchmark for model performance in clinical risk prediction. We illustrate this by implementing a superlearner to predict liver fibrosis in patients with non-alcoholic fatty liver disease (NAFLD).

**Methods:** We trained a superlearner based on 23 demographic and clinical variables, with the goal of predicting stage 2 or higher liver fibrosis. The superlearner was trained on data from the Non-alcoholic steatohepatitis – clinical research network observational study (NASH-CRN, n=648), and validated using data from participants in a randomized trial for NASH (‘FLINT’ trial, n=270) and data from examinees with NAFLD who participated in the National Health and Nutrition Examination Survey (NHANES, n=1244). We compared the performance of the superlearner with existing models, including FIB-4, NFS, Forns, APRI, BARD and SAFE.

**Results:** In the FLINT and NHANES validation sets, the superlearner (derived from 12 base models) discriminates patients with significant fibrosis from those without well, with AUCs of 0.79 (95% CI: 0.73-0.84) and 0.74 (95% CI: 0.68-0.79). Among the existing scores considered, the SAFE score performed similarly to the superlearner, and the superlearner and SAFE scores outperformed FIB-4, APRI, Forns, and BARD scores in the validation datasets. A superlearner model derived from 12 base models performed as well as one derived from 90 base models.

**Conclusions:** The superlearner, thought of as the “best-in-class” ML prediction, performed better than most existing models commonly used in practice in detecting fibrotic NASH. The superlearner can be used to benchmark the performance of conventional clinical risk prediction models.

## Introduction

Regression models remain the workhorse of clinical risk prediction. For example, all recently-developed non-invasive tools developed to predict significant liver fibrosis in the past decade, rely on logistic regression models^1–4^. Advantages of regression models include their ease of development and implementation, transparency and interpretability, and widespread understanding by clinicians. Once a regression model for risk prediction is developed, it must be tested across independent and diverse cohorts to ensure appropriate performance. Even in the best-case scenario, when a regression model developed in one cohort performs well in others, lingering questions remain: is this the best model that can be developed for its specific purpose, balancing prediction performance with the aforementioned merits of regression models? More specifically, one may ask: (1) how would the regression model compare to more complex machine learning approaches? and (2) would the inclusion of additional features/covariates improve the model performance?

In this paper, we illustrate the use of an ensemble machine learning algorithm, the “superlearner”, to benchmark the performance of clinical risk prediction algorithms, especially those based on simple regression techniques. Ensemble machine learning algorithms combine predictions from multiple models into a single prediction, and are particularly useful because it is oftentimes difficult for researchers to select a single model with optimal performance *a priori*. Even after systematically assessing the performance of various models (or “learners”), it can still be challenging to select the best performing one with confidence. The superlearner algorithm integrates information across numerous base learners by using cross-validation to identify an optimal combination of their predictions^5–7^. By borrowing strength across many learners, the superlearner may better describe complexities in the underlying data, and may produce a more accurate prediction. As such, the superlearner can be thought of as a “prediction ceiling”, capturing the best-possible or close-to-the-best-possible prediction given the available information used to develop the model. The superlearner can then be used to benchmark the performance of simpler risk prediction algorithms, such as those based on regression techniques.

We illustrate the superlearner in the context of developing a non-invasive risk scoring system to predict liver fibrosis in nonalcoholic fatty liver disease (NAFLD). NAFLD is the most common cause of chronic liver disease in the world, affecting over one-quarter of the population^8^. While the severity of liver disease in patients with NAFLD varies greatly, the single most important determinant of long-term outcomes in patients with NAFLD is the extent of liver fibrosis^9^. Non-invasive methods are essential as the initial screening test to identify at risk individuals for interventions. We rigorously compare the superlearner with numerous existing non-invasive fibrosis scoring systems in two independent validation datasets, including a US-representative cohort, and demonstrate how the superlearner can be used to assess the performance of simpler models.

## Methods

### Study Design

The overarching goal of this work is to develop a superlearner to predict significant fibrosis (stage 2 or higher) in patients with NAFLD and rigorously compare the resulting risk prediction with existing non-invasive fibrosis scoring systems in two independent validation datasets, including a US-representative cohort.

The training data set is comprised of patients enrolled in the observational cohort of the non-alcoholic steatohepatitis clinical research network (NASH CRN)^10^. The first testing data set for model evaluation included subjects who participated in the Farnesoid X nuclear receptor ligand obeticholic acid for non-cirrhotic NASH (FLINT) clinical trial^11^. The second testing data set consisted of examinees who participated in the National Health and Nutrition Examination Survey from 2017 to March 2020 Pre-Pandemic period and were found to have NAFLD^12, 13^.

In selecting these data sets, our strategy was to develop and validate our model in subjects in a wide spectrum of NAFLD, including those undergoing evaluation for NAFLD in a specialty setting (NASH CRN cohort), rigorously-selected participants in a randomized controlled trial for NASH (FLINT), and subjects in a cross-sectional study sample designed to be representative of the adult US population.

The training dataset consisted of biopsy-proven NAFLD patients of NASH-CRN, the details of which are published^10^. Briefly, the study included the entire spectrum of NAFLD patients, ranging from simple steatosis to cirrhosis, and excluded patients with other forms of chronic liver disease, such as viral hepatitis or alcohol-associated liver disease. From the study data, we selected patients with available liver biopsy data from within six months of baseline data collection.

The details of the FLINT trial have also been described in detail^11^. Briefly, the trial enrolled both diabetic and nondiabetic patients with histologically-confirmed NASH regardless of fibrosis stage, and randomized them between obeticholic acid and placebo. From these study participants, individuals without complete data necessary for the analysis (e.g., liver fibrosis stage) were excluded from our analysis. Although this trial was conducted by NASH CRN, there was no overlap between subjects of the observational cohort included in the model training and the FLINT trial participants.

NHANES is a federal program conducted by National Center for Health Statistics (NCHS) to determine the health and nutritional status of the US population, employing multistage, stratified probability samples designed to be representative of non-institutionalized civilians^13^. Since 1999, NHANES has collected and released data in 2-year cycles. NHANES data collection for 2019-2020 was suspended in March of 2020 due to COVID-19 pandemic rendering this sample not nationally representative; therefore this partial sample was combined with the 2017-2018 cycle. Sample weights and variance units were constructed and provided for this unique NHANES cycle by NCHS to produce nationally representative estimates from this 3.2-year period sample. In addition to a wide array of existing variables from questionnaires, physical examinations and laboratory data, transient elastography was added to NHANES beginning in 2017. Thus, from the 2017 to March 2020 Pre-Pandemic data set, we selected adult (>=18 years) examinees with evidence of hepatic steatosis, as defined by a controlled attenuation parameter (CAP) score of 274 or higher^14^.

From the NHANES set, subjects were excluded if there was positive viral hepatitis B or C markers and significant alcohol use, as defined by an average of more than 2 drinks per day for men or more than 1 drink per day for women; those who had 4-5 drinks on an occasion at least 5 times in the past 30 days; and those who 4 or more drinks per day at least 3-4 times a week in the past year. Individuals with incomplete elastography exam status, missing labs, or missing alcohol information were also excluded.

In the NHANES set, subjects were determined to be diabetic if they were told they had diabetes; take insulin or oral hypoglycemic agents; or had glycohemoglobin greater than 6.5%. Hypertension was present if they were told they had hypertension or high blood pressure; had a non-missing age at which they were told they had hypertension; take prescribed medication for hypertension or high blood pressure; had systolic blood pressure recorded above 140 mmHg; or had diastolic blood pressure recorded above 90 mmHg.

## Predictor and Outcome Variables

*Outcomes:* In the NASH-CRN and FLINT datasets, stage 2 or higher biopsy-proven fibrosis was the binary outcome of interest. In the NHANES-NAFLD cohort, biopsy-based fibrosis data was not available, and thus we used liver stiffness measurements (LSM) of greater than 8 KPa as the closest surrogate for the outcome interest^14^.

*Covariates:* To train the superlearner, we used data on 23 clinical, demographic and laboratory measurements common to all three datasets: age, sex, race (white v. other), Hispanic ethnicity, presence/absence of type II diabetes status, presence/absence of hypertension, albumin, alanine aminotransferase (ALT), alkaline phosphatase, aspartate aminotransferase (AST), body mass index (BMI), gamma-glutamyl transferase (GGT), globulin, fasting glucose, HDL cholesterol, hematocrit, hemoglobin A1C, LDL cholesterol, platelets, total bilirubin, total cholesterol, triglycerides, and white blood cell count.

## Statistical Analysis

The superlearner is an ensemble method that combines many “base” models into a single “super” model by using cross-validation (CV) to identify an optimal weighted combination of the base models. These base models may range from the familiar (generalized) linear regression to more sophisticated approaches like regularized regression (e.g. lasso), tree-based methods (e.g. random forest), and other sophisticated machine learning algorithms such as neural networks and support vector machines^7, 15^.

The superlearner algorithm is depicted graphically in **Figure 1**, with *V*=10 folds used for cross-validation and *M*=12 base models, as used in this study. First, we randomly split the training dataset into *V* non-overlapping and approximately equally-sized folds. For a given fold, we train all *M* base models on the other *V*-1 folds. Then, we use each of the base models to predict the outcome value for each observation in the selected fold. In the example depicted in **Figure 1**^16^, we fit each base model to the data in Folds 2-10 (holding out Fold 1); Folds 1, 3-10 (holding out Fold 2); and so on until there are 10 fits for each base model. We predict the outcomes in Fold 1 using the base models fit to Folds 2-10; predict the outcomes in Fold 2 using the base models fit to Folds 1, 3-10; and so on until we have a vector of predicted outcome values for each of the 12 base models. Note that these predictions are always on data folds not used to train the models.

**Figure 1:**
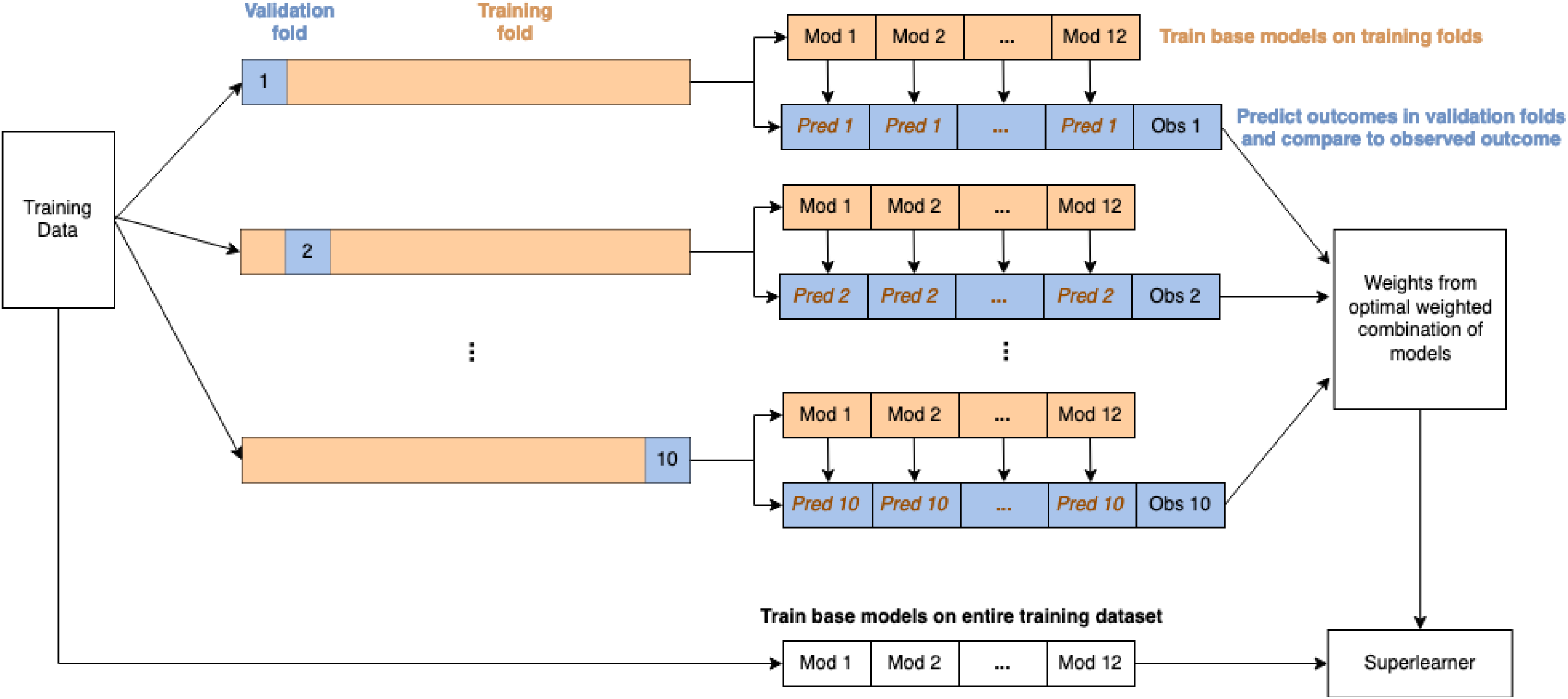
Superlearner algorithm construction illustrated with 10-fold cross-validation and 12 base models (a different number of folds or models may be used). Loosely adapted from (16).

Now we want to find the weighted combination of the CV predicted values for each base model that minimizes a pre-specified loss function, such as the mean squared error (MSE). For the MSE, the optimal weights are equivalent to the coefficient estimates from regressing the *M* sets of predicted values against the true outcome (linear regression for continuous outcomes, logistic regression for binary outcomes). Other loss functions may be used, and for ease of interpretation, the weights are generally restricted to be positive values that sum to 1. Base models that give good predictions will be assigned larger weights, while the weights associated with models that perform poorly will approach zero. Finally, we re-fit each base model to the entire training dataset. To predict new values using the superlearner, predictions from these base model fits are combined according to the weights estimated from cross-validation.

The superlearner has been shown to perform asymptotically as well as the “true” best weighted combination of the specified base learners, assuming that none of the individual prediction models correctly match the real data-generating process^5, 15^. Therefore, adding more base learners should always improve the performance of the superlearner in theory, but can be impractical beyond a certain extent, due to excessive burden in computation an collecting relevant prediction features.

Cross-validation techniques can be and oftentimes are used to tune hyperparameters of the individual base learners. Here, we are not concerned with trying to choose the best base model or model parameter(s), so it is not necessary to optimize base model hyperparameters, using CV or otherwise. Within the same superlearner, it is possible to include multiple instances of the same model with different parameter values, as we have done in this analysis.

We fit a total of six superlearner models using the SuperLearner R package^11^. First, we constructed a superlearner using all available predictor variables and the following 12 base models: Bayesian generalized linear model (bayesglm^17^), multivariate adaptive regression splines (earth^19^), generalized additive model (gam^18^), generalized boosted model (gbm^19^), generalized linear model (glm), regularized generalized linear model (glmnet^20^), bagging trees (ipredbagg^23^), neural network (nnet^21^), multivariate adaptive polynomial spline regression (polymars^22^), random forest (randomForest^23^), recursive partitioning tree (rpart^24^), and support vector machine (svm^25^). Default tuning parameters were used for all 12 base models. We then re-fit this superlearner to training data where all continuous predictors have been log transformed, and again to both the untransformed and log-transformed data. The other three superlearners were constructed from 90 base models, fit to the untransformed, log-transformed, and untransformed + log-transformed training data. These 90 base models are instances of the 12 model types used in the superlearners with 12 base models, but with varying model parameters (**Table S2**). For all superlearners, we used 10-fold CV and maximized the area under the receiver operating characteristic (ROC) curve to obtain the model weights.

For each validation cohort, we applied the six superlearner models and six additional risk scores including the AST to Platelet Ratio Index (APRI)^26, 27^, the BARD score^4^, the Fibrosis-4 (FIB-4) score^1^, the Forns index^28^, the NAFLD fibrosis score (NFS)^2^, and the Steatosis-Associated Fibrosis Estimator (SAFE) score^3^ for identifying significant liver fibrosis. In evaluating the performance of the six scores, and the superlearners in each validation cohort, we used ROC curves and area under the ROC curve (AUC) as summary measures of discriminative ability. For the NHANES-NAFLD dataset, we used sampling weights to obtain weighted AUCs and ROC curves^29^. 95% percentile bootstrap confidence intervals were computed for the AUCs using 1000 bootstrap replicates. In addition, descriptive statistics reported for the NHANES-NAFLD data account for the sample weights, clusters, and strata in its complex survey design^30, 31^.

The superlearner is a “black box” approach, which makes interpretation of its results difficult. To allow for interpretation and visualization, we plotted standardized mean differences within quartiles of the predicted superlearner and SAFE scores, as the latter turned out to perform the best among all 12 models tested. That is, for all continuous covariates considered in the validation cohorts, we standardized the variables by subtracting the mean and dividing by the standard deviation. Binary covariates were not standardized. Then, we grouped the data by quartiles defined using the predicted superlearner and SAFE scores. Finally, we calculated the mean of each predictor variable within quartiles, which can be interpreted as a standardized mean for continuous covariates and an estimated proportion for binary covariates. Confidence intervals were computed using a normal approximation (Wald confidence interval for binary variables). For the NHANES-NAFLD cohort, we used weighted means and quartiles, as well as standard deviations and standard errors that appropriately account for the complex survey design. This process allows us to characterize those who are predicted to be at high risk of significant fibrosis, compared to those with low or moderate predicted risk.

*Supplementary analyses:* Liver fibrosis stages range from 0 (no fibrosis) to 4 (cirrhosis). We selected our primary analysis calibrated for stage 2 fibrosis, which is the inflection point beyond which the risk of future morbidity and mortality rises significantly. We performed a sensitivity analysis fitting a superlearner model to predict stage 3 fibrosis (advanced fibrosis), which many of the existing models were developed to diagnose. We compared the performance of the superlearner-F3 model with existing non-invasive fibrosis scores. In the NHANES-NAFLD cohort, LSM greater than 12 KPa was used as the surrogate outcome of interest.

*Reproducibility:* All statistical analyses were performed in the R programming language^32^. R code to reproduce the figures, tables, and analysis presented in this manuscript is available at https://github.com/janewliang/NAFLD_superlearner. The official guide to the R SuperLearner package can be found at https://cran.r-project.org/web/packages/SuperLearner/vignettes/Guide-to-SuperLearner.html.

## Results

Our goal was to develop ensemble models to accurately predict significant liver fibrosis (stage II and higher) using routinely collected clinical and laboratory data using the superlearner algorithm (**Figure 1**). We trained superlearner models using data from NASH-CRN and tested these models using data from the FLINT and NHANES-NAFLD cohorts (**Figure 2**). We compared the performance of the superlearner models to six existing non-invasive fibrosis scores: APRI, BARD, FIB-4, Forns index, NFS and SAFE (**Table S1**).

**Figure 2:**
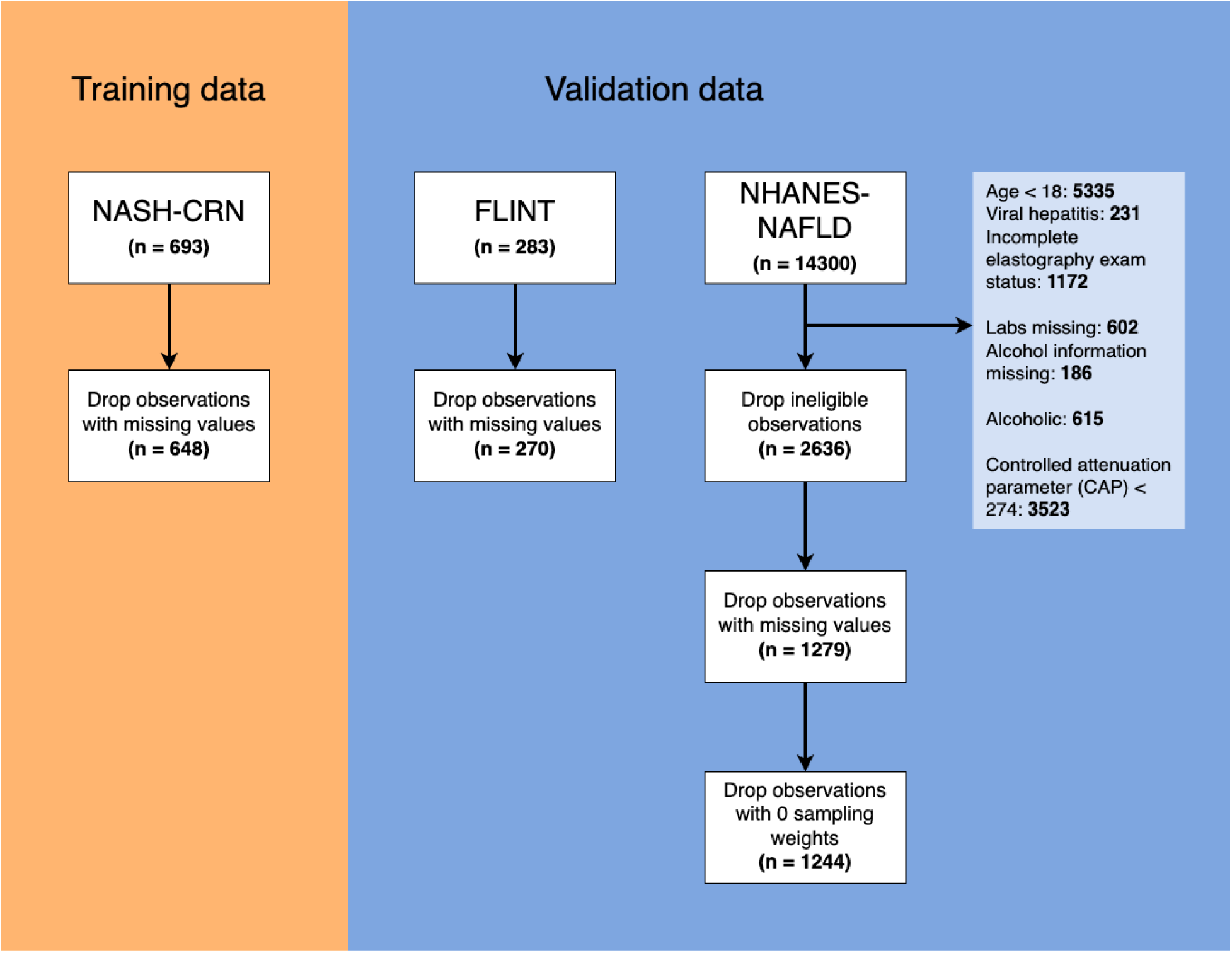
Subject selection for the training and validation cohorts used for analysis. *NHANES participants were deemed ineligible and excluded from our study if they were less than 18 years of age; had viral hepatitis; had incomplete elastography exam status, missing labs, or missing alcohol information; were alcoholic; or had controlled attenuation parameter (CAP) below 274. Alcoholic status was defined as those who had an average of more than 2 drinks per day for men or more than 1 drink per day for women; those who had 4-5 drinks on an occasion at least 5 times in the past 30 days; and those who 4 or more drinks per day at least 3-4 times a week in the past year.

In total, 23 clinical/demographic and laboratory parameters were common to the three datasets studied and were used to train the superlearner; in contrast, existing non-invasive fibrosis scores use between two and seven variables for fibrosis prediction (**Table S1**). Summary statistics for the 23 variables for in each cohort are provided in **Table 1**. Briefly, the NHANES-NAFLD cohort had lower median AST and ALT levels compared to the NASH-CRN and FLINT cohorts, and fewer patients had type II diabetes than in the FLINT cohort. Importantly, the prevalence of significant fibrosis (defined as stage II on biopsy in NASH-CRN and FLINT cohorts and LSM>8KPa in the NHANES-NAFLD cohort) was lower in the NHANES-NAFLD cohort than in the NASH-CRN and FLINT cohorts (15% v. 45-59%).

**Table 1.** Summary of laboratory and clinical characteristics in each cohort. The median (interquartile range) is reported for continuous variables and the mean (standard deviation) for binary variables. The descriptive statistics for NHANES-NAFLD adjust for survey sampling.

|  | **NASH-CRN**<br>**(Training)** | **FLINT**<br>**(Testing #1)** | **NHANES-**<br>**NAFLD**<br>**(Testing #2)** |
| --- | --- | --- | --- |
| **Albumin (g/dL)** | 4.2 (0.5) | 4.3 (0.6) | 4 (0.4) |
| **Alanine aminotransferase (U/L)** | 64.5 (54) | 68 (61.5) | 21 (15) |
| **Alkaline phosphatase (U/L)** | 80 (36) | 76.5 (34.75) | 74 (26) |
| **Aspartate aminotransferase (U/L)** | 46 (34) | 51 (39.5) | 19 (8) |
| **Body mass index (kg/m<sup>2</sup>)** | 33.66 (8.6) | 33.58 (7.29) | 32.1 (8.7) |
| **Gamma-glutamyl transferase (U/L)** | 49 (54) | 48.5 (58) | 24 (16) |
| **Globulin (g/dL)** | 3 (0.7) | 3 (0.6) | 3 (0.5) |
| **Glucose (mg/dL)** | 95 (24) | 103.5 (31) | 109 (21) |
| **HDL cholesterol (mg/dL)** | 42 (14) | 42 (14) | 45 (16) |
| **Hematocrit (%)** | 42.4 (4.8) | 41.05 (4.88) | 43 (5.3) |
| **Hemoglobin A1C (%)** | 5.7 (0.9) | 6.2 (1.2) | 5.7 (0.7) |
| **Hypertension (yes/no)** | 0.44 (0.5) | 0.61 (0.49) | 0.54 (0.5) |
| **LDL cholesterol (mg/dL)** | 119 (48) | 108.5 (52.75) | 112 (48) |
| **Platelets (1000/mm<sup>3</sup>)** | 245 (80.5) | 232.5 (74.5) | 237 (77) |
| **Total bilirubin (mg/dL)** | 0.7 (0.4) | 0.6 (0.4) | 0.4 (0.3) |
| **Total cholesterol (mg/dL)** | 193 (51.25) | 185 (63.25) | 183 (52) |
| **Triglycerides (mg/dL)** | 149 (99) | 154 (92.5) | 116 (79) |
| **Type 2 diabetes (yes/no)** | 0.22 (0.42) | 0.49 (0.5) | 0.25 (0.43) |
| **White blood cell (1000/mm<sup>3</sup>)** | 6.7 (2.4) | 6.85 (2.7) | 7 (2.4) |
| **Age (years)** | 49 (17) | 53 (16.75) | 54 (26) |
| **Hispanic ethnicity (yes/no)** | 0.14 (0.35) | 0.16 (0.36) | 0.19 (0.4) |
| **Male gender (yes/no)** | 0.38 (0.49) | 0.34 (0.48) | 0.55 (0.5) |
| **White race (yes/no)** | 0.8 (0.4) | 0.82 (0.38) | 0.62 (0.49) |
| **Significant fibrosis (yes/no)** | 0.45 (0.5) | 0.59 (0.49) | 0.15 (0.36) |

We present results from a superlearner combining 12 base models in which we allowed continuous predictors to be on their natural scale or log-transformed, with results from other more complex superlearner models presented in the supplement. The estimated optimal associated weights underlying this superlearner model are provided in **Table 2**; ∼23% of the model weights are assigned to multivariate adaptive polynomial spline regression (polymars), and the remaining ∼77% of the model weights are spread among other base learners. As such, this superlearner represents the optimal combination of the 12 base models to predict stage II or higher fibrosis.

**Table 2.** Estimated optimal model weights for the superlearner, constructed from 12 base models, considering data on 23 clinical/demographic variables. Both untransformed and log-transformed continuous variables were considered.

|  | **Superlearner-12**<br>**(all)** |
| --- | --- |
| Bayesian generalized linear model | 0.07 |
| Multivariate adaptive regression splines | 0.029 |
| Generalized additive model | 0 |
| Generalized boosted model | 0.146 |
| Generalized linear model | 0.001 |
| Regularized generalized linear model | 0.122 |
| Bagging trees | 0.062 |
| Neural network | 0.102 |
| Multivariate adaptive polynomial spline regression | 0.227 |
| Random forest | 0.165 |
| Recursive partitioning tree | 0 |
| Support vector machine | 0.074 |

**Figures 3 and 4** demonstrate the ROC curves and AUCs for the superlearner compared to the other six existing fibrosis scores for comparison, in the two testing datasets: FLINT and NHANES-NAFLD. In both data sets, the superlearner was able to discriminate patients with significant fibrosis from those without well, with AUCs of 0.79 (95% CI: 0.73-0.84) and 0.74 (95% CI: 0.68-0.79), respectively. In the FLINT cohort, besides the superlearner, SAFE had the highest AUC, (though confidence intervals overlap with APRI, FIB-4, and NFS). A similar pattern was replicated in the NHANES-NAFLD cohort, in which the NFS score had marginally lower AUC than the superlearner and SAFE scores. The BARD, FIB-4 and Forns index scores performed noticeably worse than the superlearner, NFS and SAFE scores in the NHANES-NAFLD cohort. As the SAFE score produced a comparable AUC to that of the superlearner (**Figure 4**), in **Figure S1,** we examine their correlation and illustrate that the superlearner and SAFE scores are in close alignment (Spearman’s correlation coefficient of 0.91-0.93 in both cohorts).

**Figure 3:**
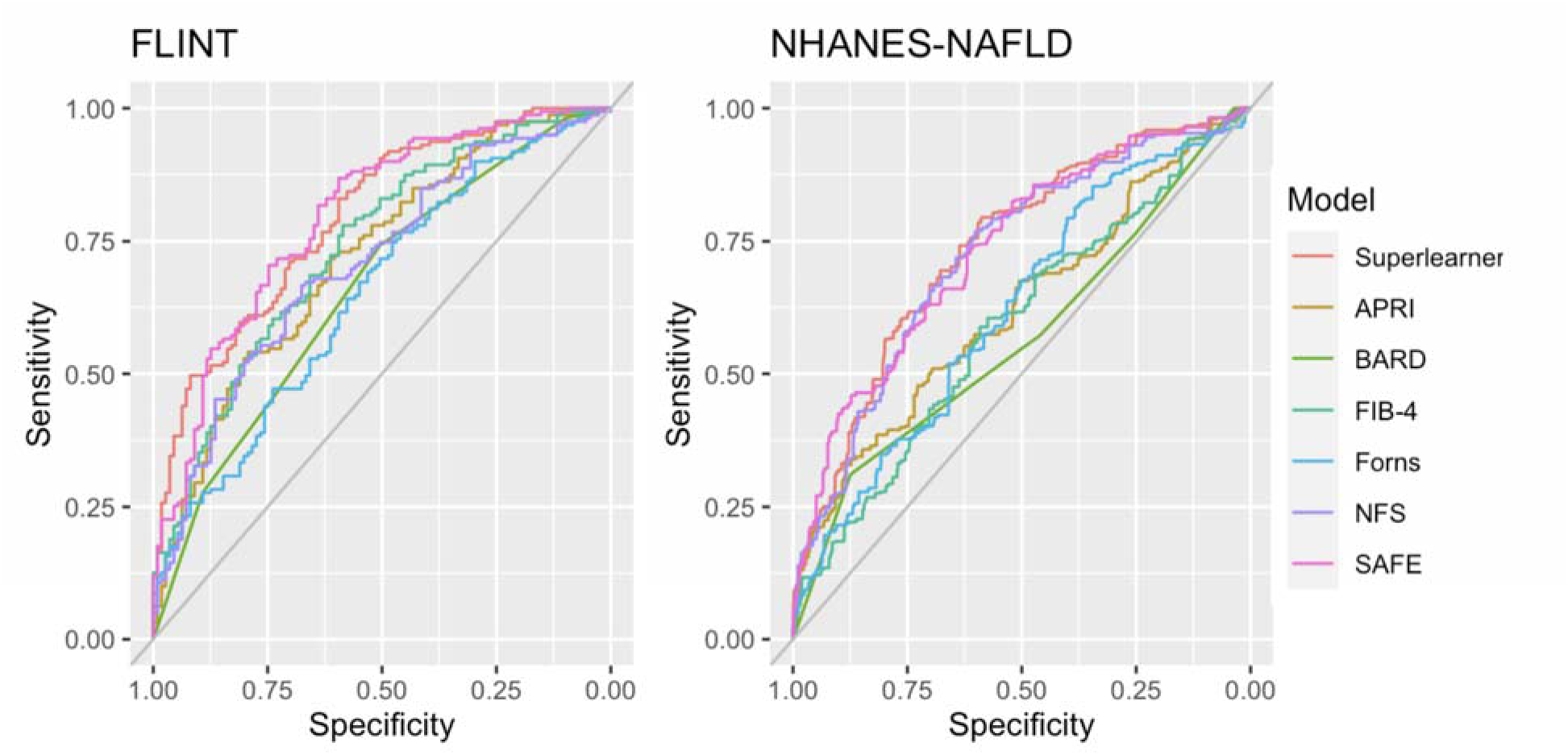
Receiver operating characteristic (ROC) curves for superlearner (based on 12 base models), APRI, BARD, FIB-4, Forns, NFS, and SAFE applied to each validation dataset. The NHANES-NAFLD ROC curves are weighted to account for survey sampling.

**Figure 4:**
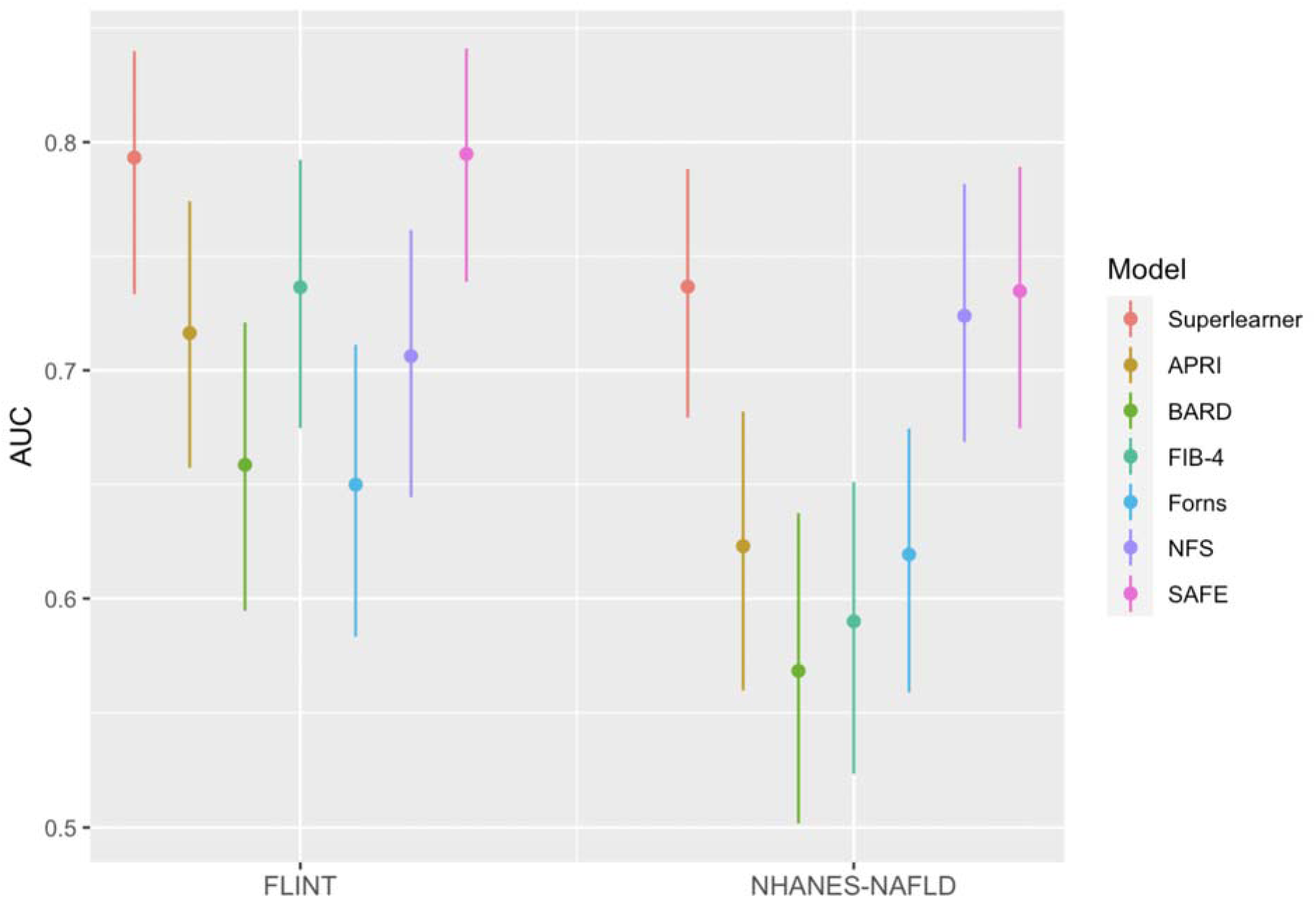
Area under the ROC curve (AUC) for superlearner (based on 12 base models), APRI, BARD, FIB-4, Forns, NFS, and SAFE applied to each validation dataset. 95% bootstrap confidence intervals are also shown. The NHANES-NAFLD AUCs are weighted to account for survey sampling.

In sensitivity analyses, we considered other variations of the superlearner using more models (ensembles of up to 90 models total, **Table S2**), but the predictive performance of all the superlearner models considered were similar (**Table S3**). While including additional base models is generally believed to improve predictive performance, we found that in this context, the superlearners with 90 base models did not consistently outperform the considerably less complex and computationally intensive superlearners with only 12 base models. The superlearners also gave similar results despite notable differences in base model weights (**Table S3**).

To better visualize the covariate profiles of patients predicted to have significant fibrosis by the superlearner with 12 base models, we plotted the standardized means and estimated proportions within predicted superlearner score quartiles for continuous and binary predictors, respectively (**Figure 5**). In both the FLINT and NHANES-NAFLD cohorts, those in the fourth quartile (predicted to be at highest risk of significant fibrosis) tended toward being older; having higher BMIs; having type 2 diabetes or hypertension; exhibiting higher levels of AST, GGT, fasting glucose, and hemoglobin A1C; and exhibiting lower levels of albumin and platelets. On average, the high-risk group in the FLINT cohort had higher ALT, ALP, and globulin levels, as well as a lower standardized mean for hematocrit. In the NHANES-NAFLD cohort, the high-risk group was associated with lower average total and LDL cholesterol levels.

**Figure 5:**
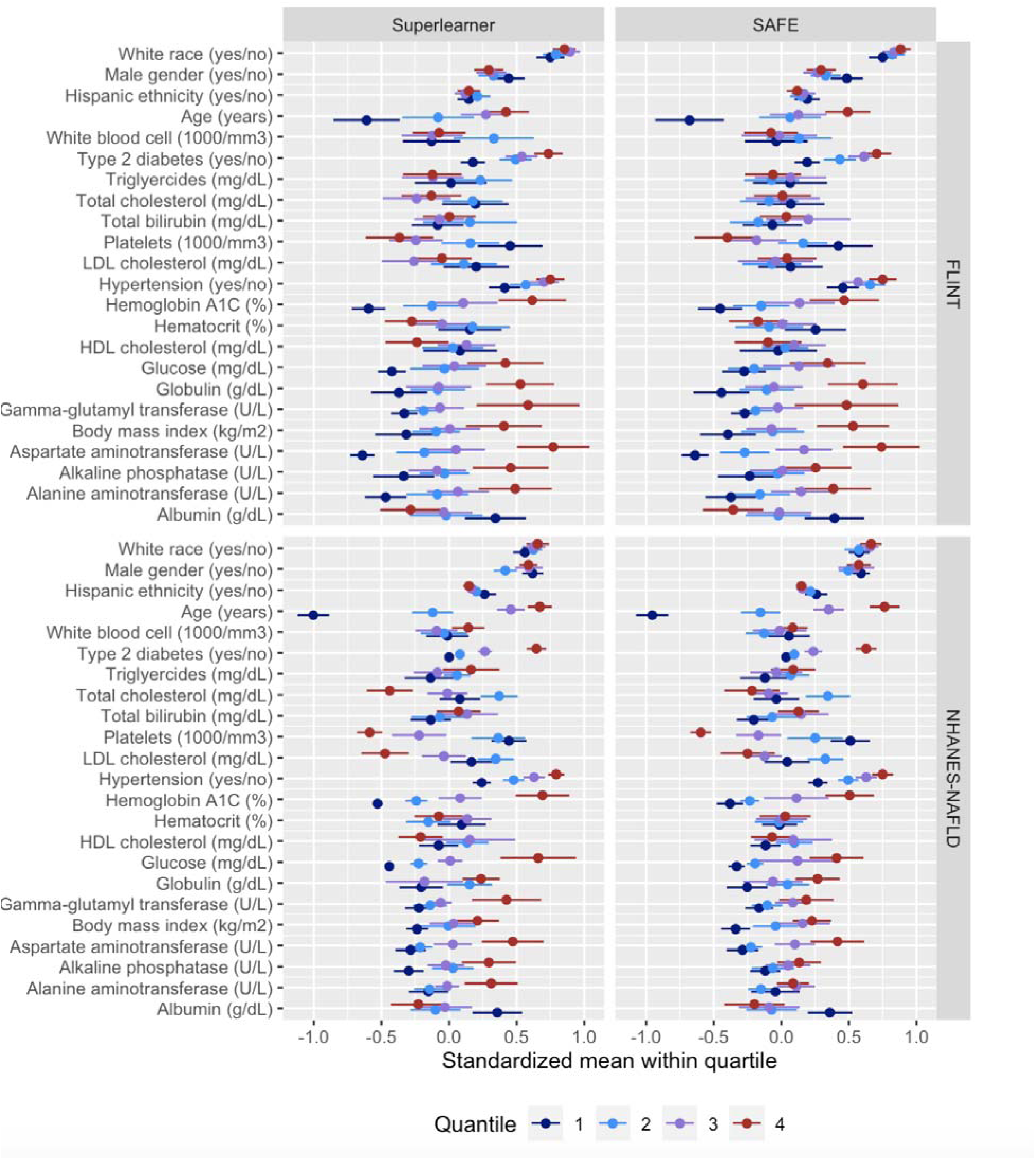
Standardized mean within quartile (with 95% normal approximation confidence intervals) for each continuous predictor and the estimated proportion within quartile (with 95% Wald confidence intervals) for each binary predictor in the FLINT and NHANES-NAFLD cohorts. Quartiles were defined based on the superlearner (12 base models) or SAFE scores. For the NHANES-NAFLD dataset, the calculation of means, standard deviations, quartiles, and confidence intervals incorporates its complex survey design.

In supplementary analyses, we explored how each of the non-invasive fibrosis scores, including the superlearner, correlate with liver stiffness measures (LSM) in the NHANES-NAFLD dataset (**Figure S2**). The Spearman’s correlation (adjusted for survey sampling) between each of the predicted scores and LSM is slightly positive for superlearner, NFS, and SAFE, and lower for the other scores. The correlation is essentially zero for FIB-4 and BARD, which also have the lowest AUCs. We also compared the performance of each of the non-invasive fibrosis scores to the FibroScan-AST (FAST) score and the Agile3+ and Agile4 scores in the NHANES-NAFLD cohort. Overall, the superlearner had significant correlation with these scores (**Figure S3-S5**)

Finally, we fit a superlearner model to predict stage III fibrosis (F3) and higher (**Figure S6**). We found that the superlearner-F3 had excellent performance in both the FLINT and the NHANES cohorts (AUCs: 0.78-0.79), comparable to the SAFE score. In contrast, other non-invasive fibrosis predictors had worse performance (AUCs: 0.6-0.7; **Figure S6**).

## Discussion

In this paper, we implement an ensemble machine learning algorithm to benchmark the performance of simpler risk prediction models, using the prediction of significant liver fibrosis in patients with NAFLD as an example. The superlearner model was trained to optimally combine the results of 12 (and up to 90) individual statistical and machine learning algorithms, using 23 laboratory and demographic variables. The superlearner competently recognized significant fibrosis (stage II and higher) in two independent testing datasets, and performed better than existing models commonly used in practice, namely FIB-4 and NFS scores.

The appeal of an ensemble superlearner model is that it allows researchers to combine predictions from many conventional and novel machine learning algorithms and avoid committing to a single, possibly poor-performing, algorithm *a priori*^7^. The superlearner has been shown to perform asymptotically as well as the best weighted combination of the specified base learners^15^. As more prediction features and more complex machine learning techniques become available, there is a need to explore whether modern machine learning methods can tangibly improve on conventional statistical approaches to prediction by leveraging high dimensional data and modeling potentially complex relationships between predictors and liver fibrosis. Each existing non-invasive risk score represents a single algorithm trained on a single dataset. In general, researchers and clinicians cannot identify one algorithm as “the best” *a priori*.

Ensemble machine learning approaches offer the opportunity to achieve better predictive performance across diverse datasets by combining potentially diverse models into a single prediction, whose performance tends to be more robust to individual learners and close to the “performance ceiling”. Therefore, the developed superlearner may play two important roles in practice: (1) it can be used to replace an individual risk prediction algorithm for its superior performance; and (2) it can serve as a benchmark to determine if the existing risk prediction algorithm is already sufficiently accurate. We note that depending on the context, even if the superlearner outperforms individual learners, one may still prefer an individual learner due to the computational burden of the superlearner and its lack of interpretability. However, by comparing the performance of an individual learner to the superlearner, we can understand whether the individual learner has large room for improvement.

We focused on liver fibrosis in NAFLD as an example, since developing clinically useful non-invasive risk scores for liver fibrosis has been an important yet challenging task in clinical medicine for the past decade. Many models have been created, analyzing and synthesizing information derived from different clinical, laboratory and demographic variables to estimate liver fibrosis. Our work demonstrates that superlearner models that consider many more variables than the other existing non-invasive predictors (23 v. 2-7 variables), and are of enormous complexity (combining information from up to 90 models, for example), outperform some but not all existing non-invasive predictors. In particular, the SAFE score, based on a multivariable logistic regression model using seven variables, achieves similar performance to the superlearner for predicting both F2 and F3 or higher fibrosis in the two testing data sets studied. This finding illustrates that even when the superlearner fails to outperform a simpler model, it can be used as a benchmark to assess whether the simpler model achieves the prediction ceiling given a particular training dataset. As such, the superlearner approach, which requires minimal human input besides *a priori* selection of the base models, can be used as a robustness-check for other models to be compared against. In this specific context, our results lend credibility to the SAFE score, despite its substantially simpler form.

Several limitations must be mentioned. First, the superlearner algorithm, including choice of all base learners and tuning parameters, should be specified *a priori*, as rerunning the algorithm after adjusting parameters can lead to overfitting. Second, in the context presented in this paper, the superlearner is focused on prediction, where the goal is to predict the probability of significant liver fibrosis conditional on 23 observed covariates. The superlearner does not produce an interpretable equation that communicates the *effects* of each covariate on the probability of the outcome, though an equation/calculator to allow for implementation can be produced. To address this limitation, we present an approach to visualize the covariate profiles of the patients predicted by the superlearner to have significant fibrosis (**Figure 5**); this visualization tool gives us insight about the characteristics of the patients identified by the algorithm. The third limitation is the computational cost of fitting the superlearner. When nested cross-validation is unnecessary (e.g. in the setting where there are multiple independent training/testing datasets), the computational burden of fitting a superlearner, even one with as many as 90 models, is modest. To encourage use of the superlearner algorithm in clinical risk prediction, we have made our code publicly available.

In conclusion, using the superlearner approach, we were able to train a model to predict clinically significant liver fibrosis in patients with NAFLD. Based on the robust number of practicable variables to be included in non-invasive assessment of NAFLD, the superlearner was able to outperform existing tools that have been widely used for NAFLD and other conditions.

In our view, the superlearner represents the “best possible” prediction given a training dataset, and even in settings where it does not outperform existing simpler models, it can be used as a benchmark to assess the performance of existing clinical risk prediction models.

## Data Availability

NASH-CRN data are available from the NIH/NIDDK Central Repository. NHANES data are available from the CDC. Code to reproduce the analysis presented is available at the link provided within the article.

## Acknowledgements

*Funding information:* This work was supported by the National Institutes of Health (R01 DK-127224, P30 DK116074, KL2 TR003143, K23 AA-029197). The funding organizations played no role in the design and conduct of the study, in the collection, management, analysis, and interpretation of the data, or in the preparation, review, or approval of the manuscript.

*Conflicts of interest:* The authors have no relevant conflicts of interest to disclose.

## FIG

