## Supplementary Information for "Benchmarking clinical risk prediction algorithms with ensemble machine learning: An illustration of the superlearner algorithm for the non-invasive diagnosis of liver fibrosis in non-alcoholic fatty liver disease"

Department of Pathology

Stanford University School of Medicine

**Table S1.** Existing scores to estimate liver fibrosis and their respective components.

| Score | Variables |
| --- | --- |
| The AST to Platelet Ratio Index (**APRI**)^27,28^ | AST, platelets |
| The **BARD** score^4^ | BMI, AST, ALT, diabetes |
| The Fibrosis-4 (**FIB-4**) score^5^ | Age, AST, ALT, platelets |
| The **Forns** index^3^ | Age, platelets, gamma-glutamyl transferase, total cholesterol |
| The NAFLD fibrosis score (**NFS**)^6^ | Age, BMI, type 2 diabetes status, AST, ALT, platelets, albumin |
| The Steatosis-Associated Fibrosis Estimator (**SAFE**) score^7^ | Age, BMI, type 2 diabetes, AST, ALT, globulin, platelets |

**Table S2:** Tuning parameters used for the 90 superlearner base models, spanning 12 base model types. For a given model type, all combinations of the listed tuning parameters were used to define base model instances. If unspecified, default parameters were used; see individual R packages for further details. Let *p* indicate the number of model predictors.

| **Base model type** | **Number of instances** | **Model parameters considered** |
| --- | --- | --- |
| **bayesglm**  Bayesian generalized linear model | 1 | N/A |
| **earth**  Multivariate adaptive regression splines | 3 | **degree: 1, 2, 3**  Maximum degree of interaction |
| **gam**  Generalized additive model | 1 | N/A |
| **gbm**  Generalized boosted model | 27 | **interaction.depth: 1, 2, 3**  Maximum depth of each tree  **shrinkage: 0.1, 0.01, 0.001**  Learning rate applied to each tree  **n.minobsinnode: 1, 10, 20**  Minimum size of terminal node |
| **glm**  Generalized linear model | 1 | N/A |
| **glmnet**  Regularized generalized linear model | 3 | **alpha: 0, 0.5, 1**  Elastic-net mixing parameter |
| **ipredbagg**  Bagging trees | 9 | **minsplit: 10, 20, 30**  Minimum number of observations for split to be attempted  **cp: 0.005, 0.01, 0.05**  Complexity parameter |
| **nnet**  Neural network | 9 | **size: 2, 5, 10**  Number of units in the hidden layer  **decay: 0, 0.25, 0.5**  Weight decay |
| **polymars**  Multivariate adaptive polynomial spline regression | 9 | **gcv: 2, 4, 6**  Generalized cross validation value for choosing model  **knot.space: 2, 3, 4**  Minimum order statistics apart that two knots can be |
| **randomForest**  Random forest | 9 | **mtry = floor(sqrt(*p*)) * {0.5, 1, 2}**  Number of variables randomly sampled at each split  **nodesize: 10, 20, 50**  Minimum size of terminal node |
| **rpart**  Recursive partitioning tree | 9 | **minsplit: 10, 20, 30**  Minimum number of observations for split to be attempted  **cp: 0.005, 0.01, 0.05**  Complexity parameter |
| **svm**  Support vector machine | 9 | **gamma: (1/*p*) * {0.5, 1, 2}**  Kernel parameter  **cost: 0.5, 1, 1.5**  Cost of constraints violation |

**Table S3:** Area under the curve (AUC) for superlearners constructed from 12 or 90 base models with untransformed predictors, log predictors (log), or both untransformed and log predictors (all); APRI; BARD; FIB-4; Forns; NFS; and SAFE applied to each validation dataset. 95% bootstrap confidence intervals are also reported. The NHANES-NAFLD AUCs are weighted to account for survey sampling. The “SL-12 (all)” model (highlighted in red) is presented in the main text.

|  | **FLINT** | **NHANES-NAFLD** |
| --- | --- | --- |
| **SL-12** | 0.791 (0.734, 0.838) | 0.702 (0.644, 0.756) |
| **SL-12 (log)** | 0.783 (0.723, 0.83) | 0.738 (0.68, 0.791) |
| **SL-12 (all)** | 0.793 (0.733, 0.84) | 0.737 (0.679, 0.788) |
| **SL-90** | 0.792 (0.733, 0.84) | 0.718 (0.66, 0.772) |
| **SL-90 (log)** | 0.784 (0.723, 0.831) | 0.726 (0.668, 0.779) |
| **SL-90 (all)** | 0.801 (0.742, 0.848) | 0.716 (0.658, 0.772) |
| **APRI** | 0.716 (0.657, 0.774) | 0.623 (0.56, 0.682) |
| **BARD** | 0.659 (0.595, 0.721) | 0.568 (0.502, 0.637) |
| **FIB-4** | 0.736 (0.675, 0.792) | 0.59 (0.523, 0.651) |
| **Forns** | 0.65 (0.583, 0.711) | 0.619 (0.559, 0.674) |
| **NFS** | 0.706 (0.644, 0.762) | 0.724 (0.668, 0.782) |
| **SAFE** | **0.795 (0.739, 0.841)** | **0.735 (0.674, 0.789)** |

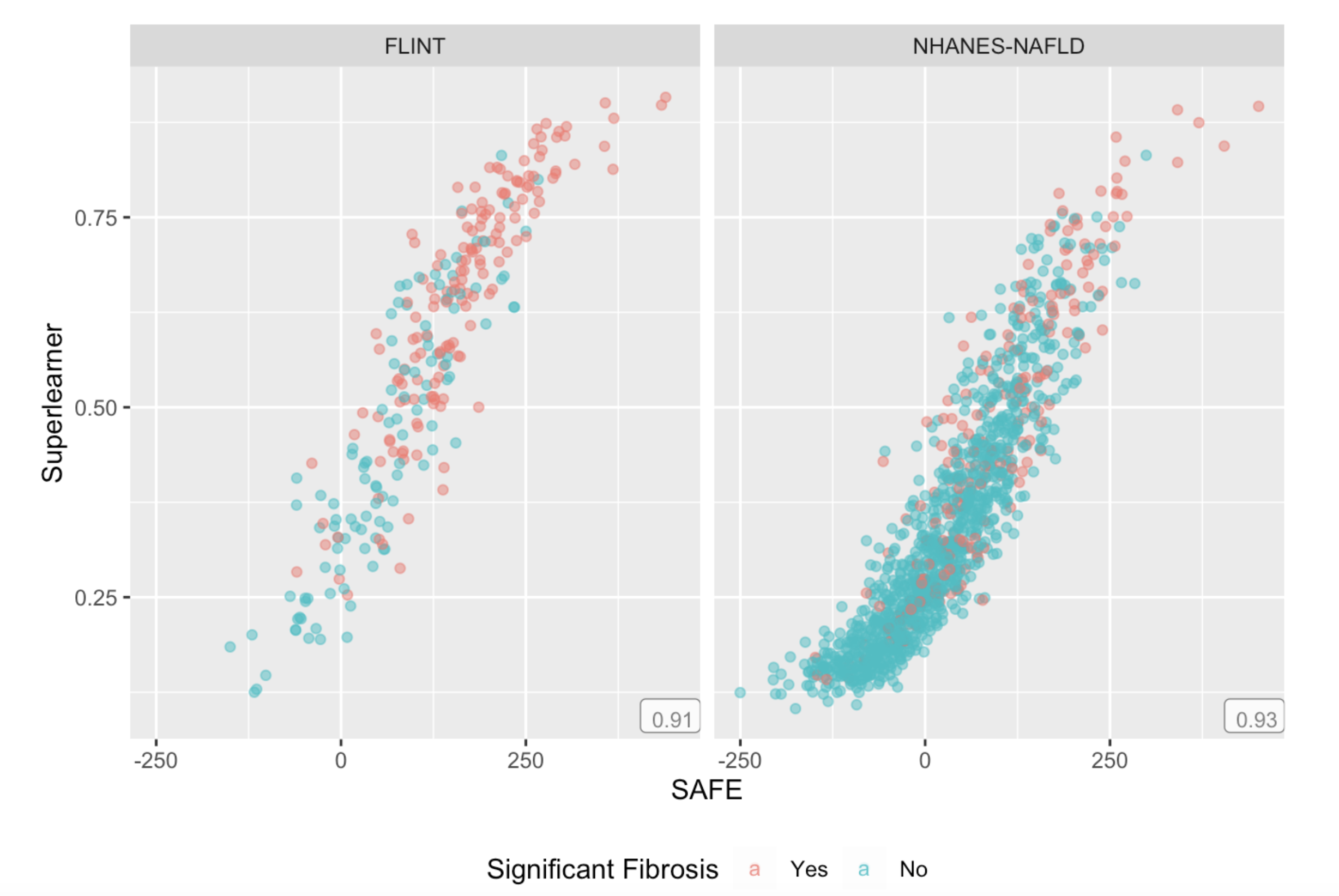

**Figure S1:** Superlearner (12 base models [“SL-12 (all)”]) v. SAFE scores predicted for the FLINT and NHANES-NAFLD validation cohorts. Individuals with significant fibrosis are indicated by point color. The Spearman's rank correlation coefficient for complex surveys is reported in the lower right of each panel.

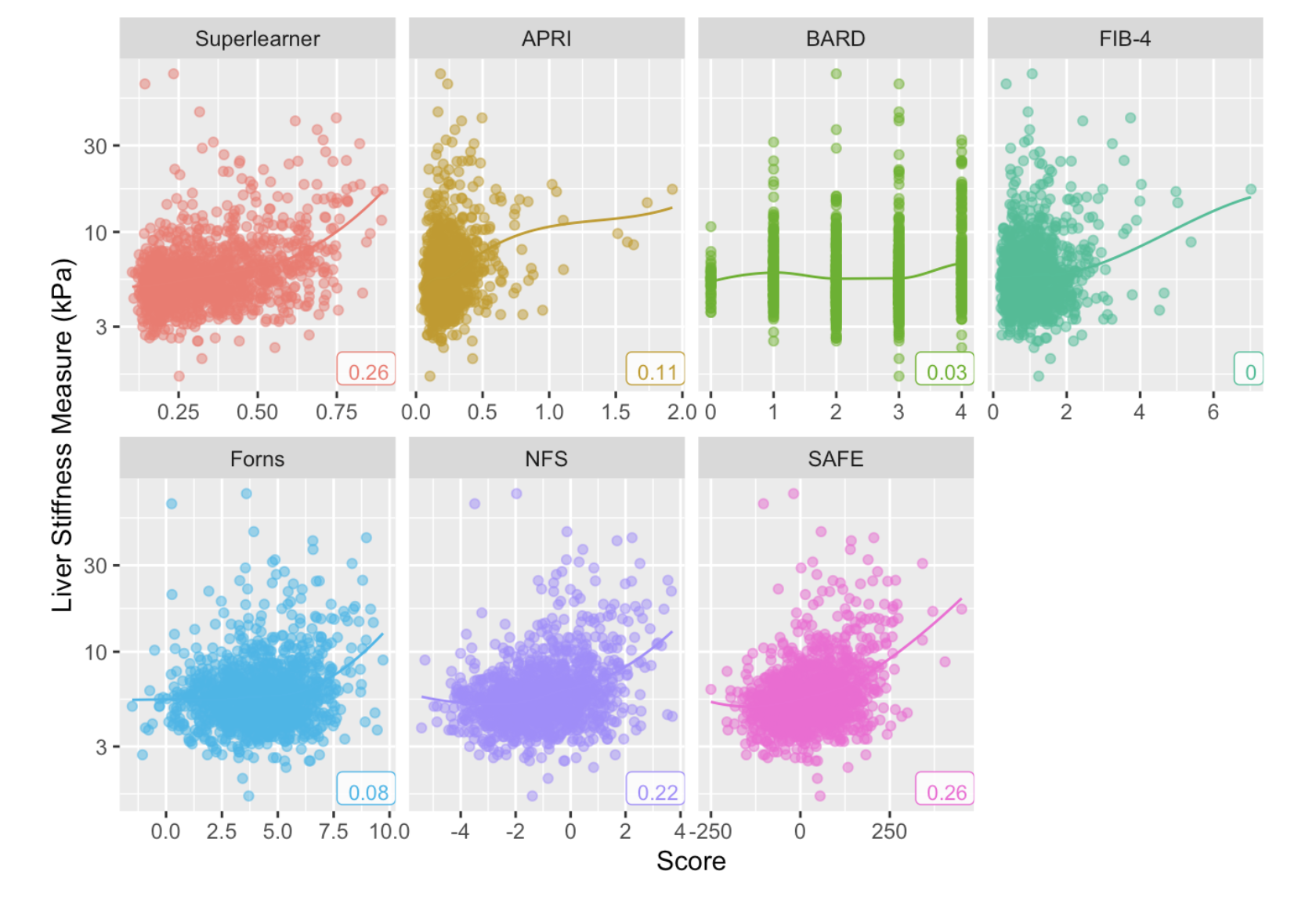

**Figure S2:** Log-transformed liver stiffness measure (kPa) plotted against predicted scores for superlearner (12 base models [“SL-12 (all)”]), APRI, BARD, FIB-4, Forns, NFS, and SAFE in the NHANES-NAFLD cohort, with loess smoothing lines. The Spearman's rank correlation coefficient for complex surveys is reported in the lower right of each panel.

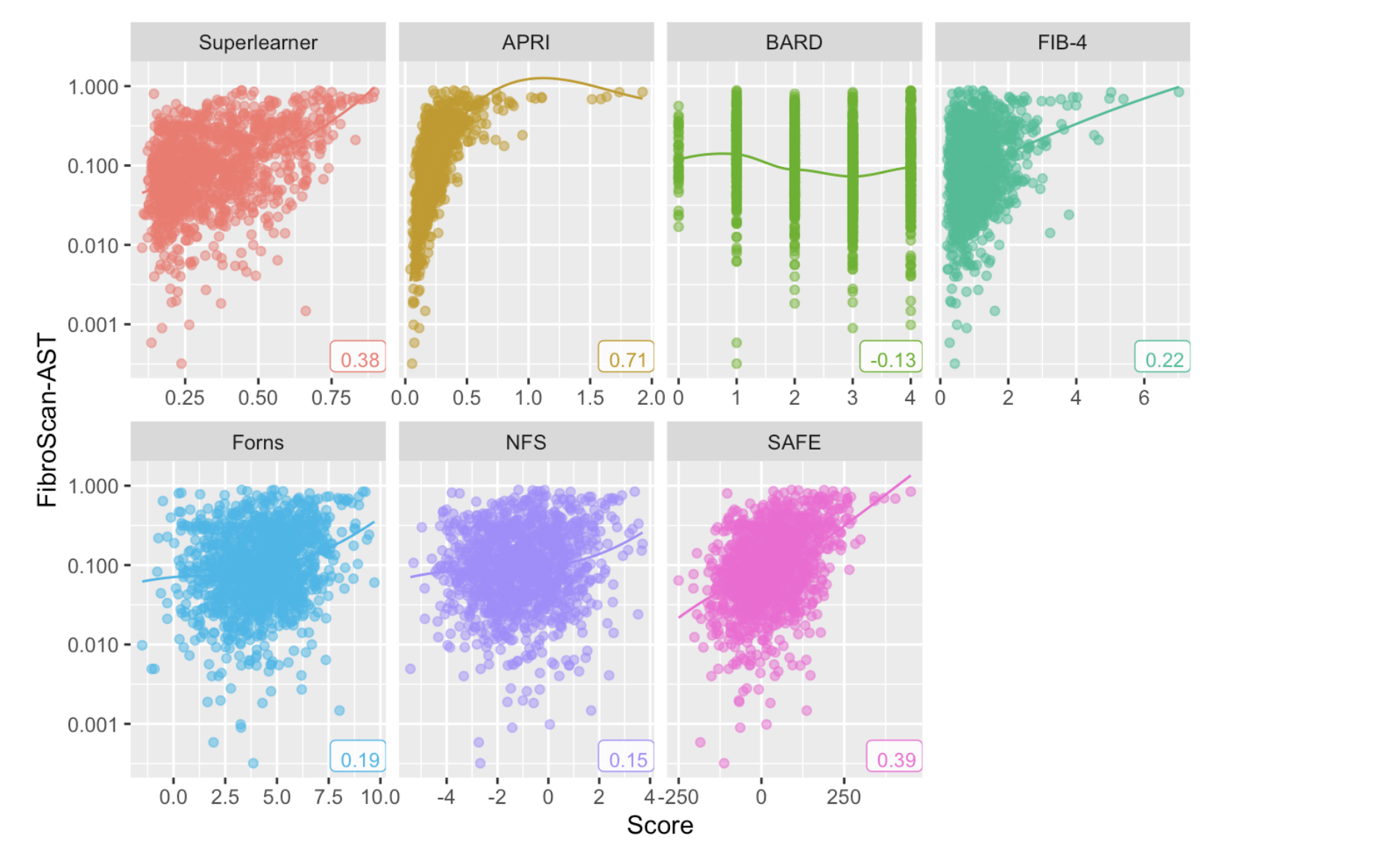

**Figure S3:** Log-transformed FibroScan-AST (FAST) score plotted against predicted scores for superlearner (12 base models [“SL-12 (all)”]), APRI, BARD, FIB-4, Forns, NFS, and SAFE in the NHANES-NAFLD cohort, with loess smoothing lines. The Spearman's rank correlation coefficient for complex surveys is reported in the lower right of each panel.

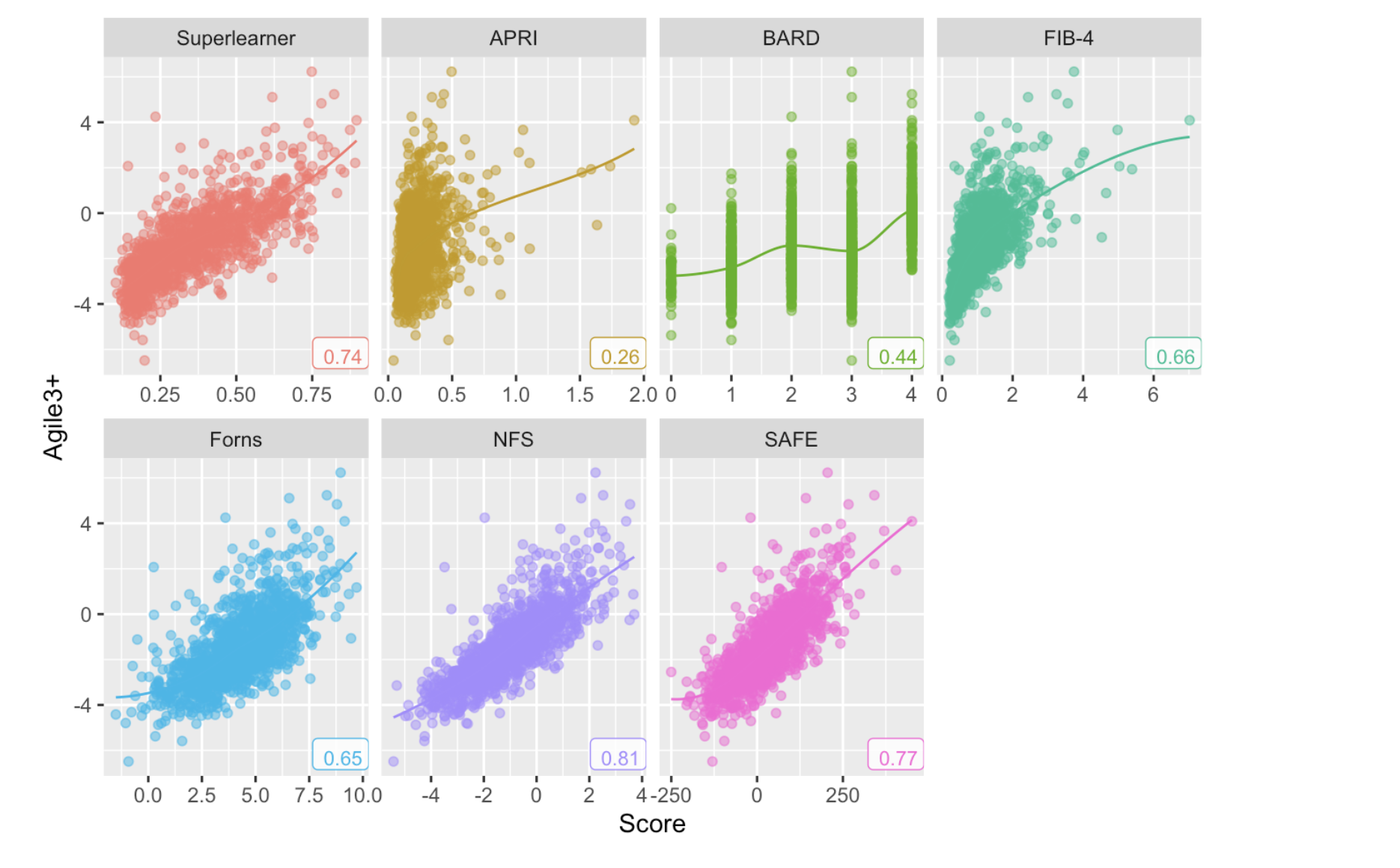

**Figure S4:** Agile3+ score plotted against predicted scores for superlearner (12 base models [“SL-12 (all)”]), APRI, BARD, FIB-4, Forns, NFS, and SAFE in the NHANES-NAFLD cohort, with loess smoothing lines. The Spearman's rank correlation coefficient for complex surveys is reported in the lower right of each panel.

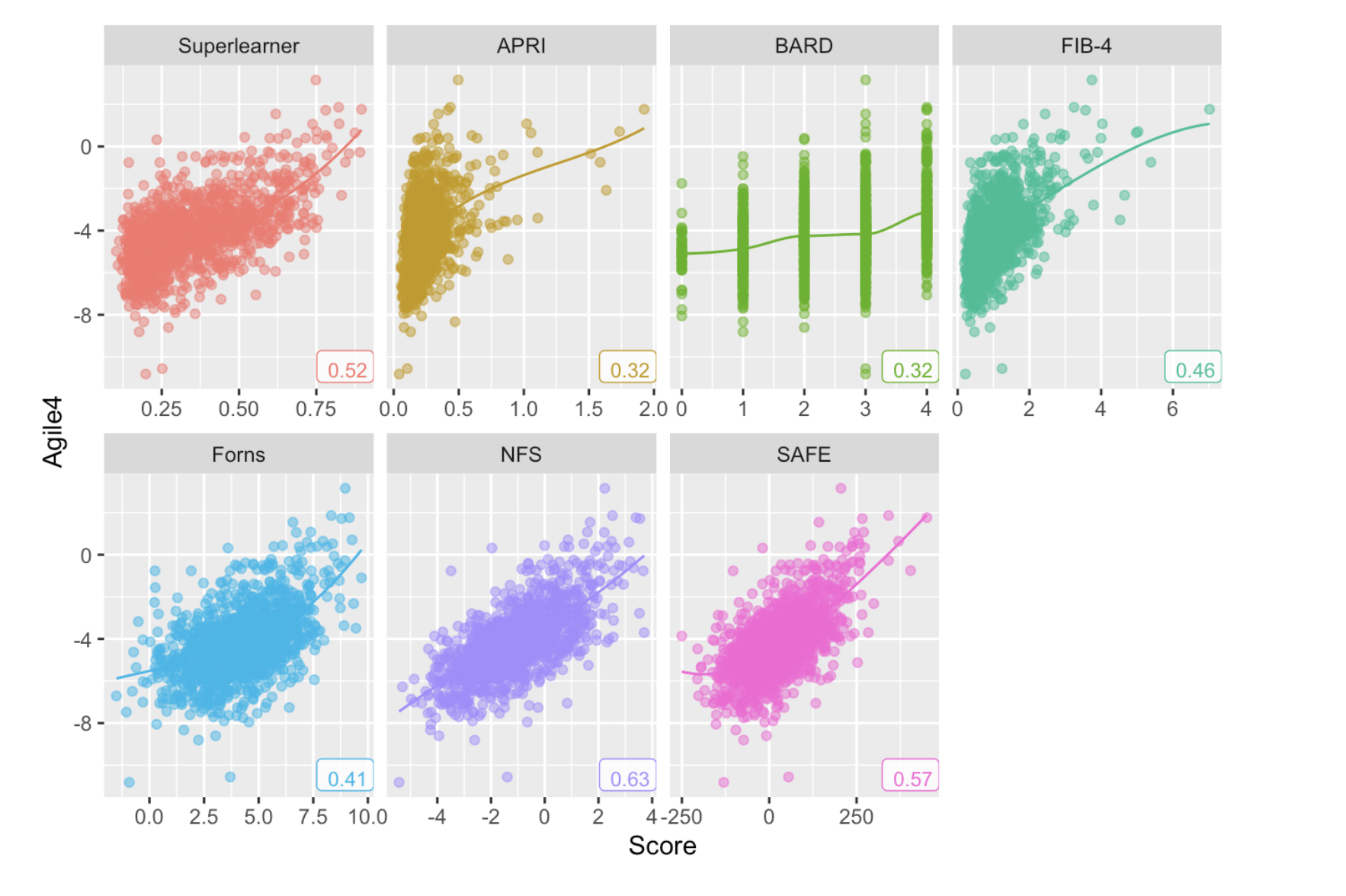

**Figure S5:** Agile4 score plotted against predicted scores for superlearner (12 base models [“SL-12 (all)”]), APRI, BARD, FIB-4, Forns, NFS, and SAFE in the NHANES-NAFLD cohort, with loess smoothing lines. The Spearman's rank correlation coefficient for complex surveys is reported in the lower right of each panel.

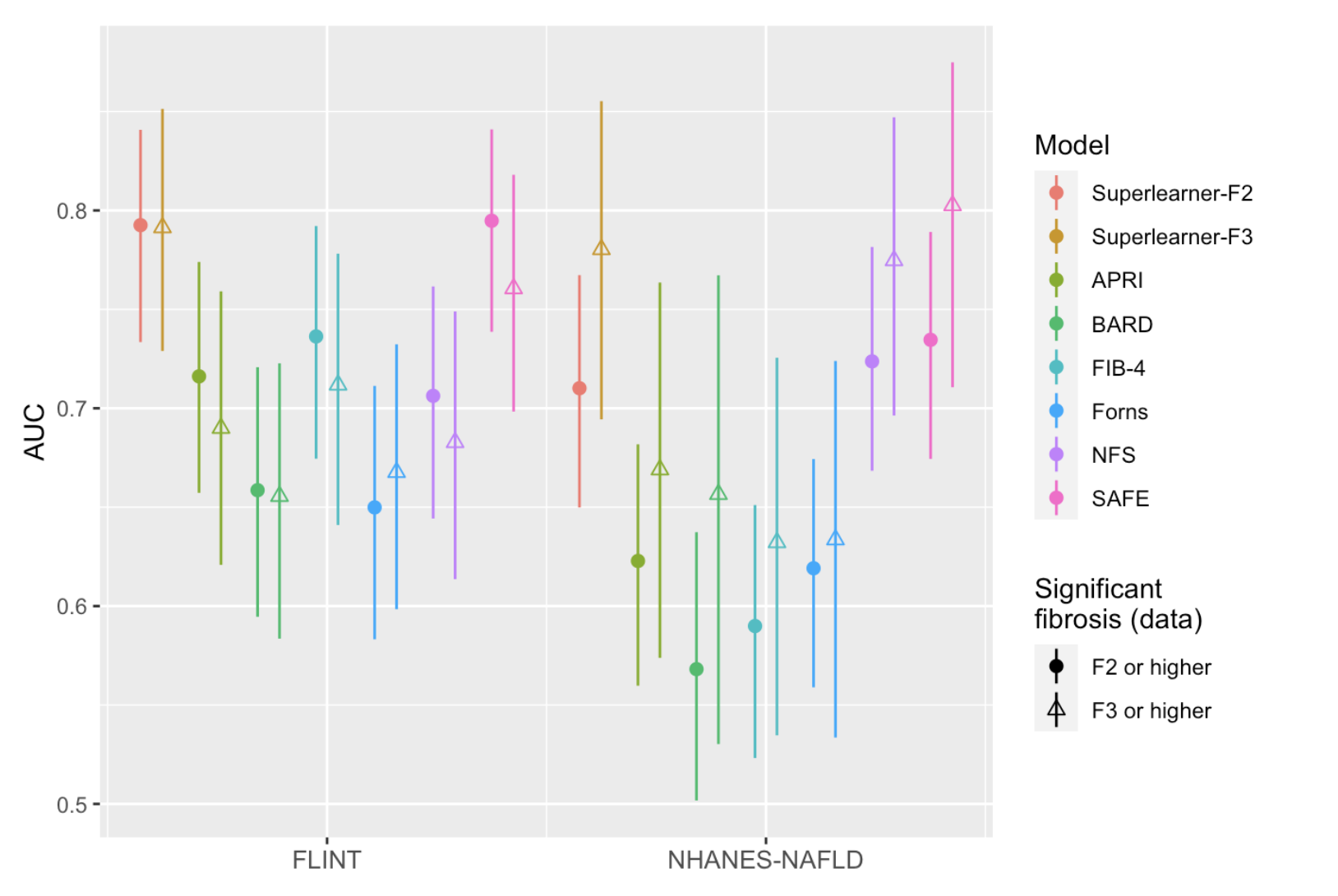

**Figure S6.** Performance of two superlearner models (one trained to predict F2 fibrosis and the other to predict F3 fibrosis, each with 12 base models) compared to other non-invasive fibrosis tests for F2 and F3 or higher fibrosis.
